# Association of mental disorders with SARS-CoV-2 infection and severe health outcomes: a nationwide cohort study

**DOI:** 10.1101/2020.08.05.20169201

**Authors:** Ha-Lim Jeon, Jun Soo Kwon, So-Hee Park, Ju-Young Shin

## Abstract

**Background:** No epidemiological data exists for the association between mental disorders and the risk of severe acute respiratory syndrome coronavirus 2 (SARS-CoV-2) infection and coronavirus disease 2019 (COVID-19) severity.

**Aims:** To evaluate the association between mental disorders and the risk of SARS-CoV-2 infection and severe outcomes following COVID-19.

**Methods:** We performed a cohort study using the Korean COVID-19 patient database based on the national health insurance data. Each patient with a mental or behavioral disorder (diagnosed during six months prior to the first SARS-CoV-2 test) was matched by age, sex, and Charlson comorbidity index with up to four patients without mental disorders. SARS-CoV-2 positivity risk and risk of death or severe events (intensive care unit admission, use of mechanical ventilation, and acute respiratory distress syndrome) post-infection were calculated using conditional logistic regression analysis.

**Results:** Among 230,565 patients tested for SARS-CoV-2, 33,653 (14.6%) had mental disorders, 928/33,653 (2.76%) tested positive, and 56/928 (6.03%) died. In multivariate analysis with the matched cohort, there was no association between mental disorders and SARS-CoV-2 positivity risk (odds ratio [OR], 1.02; 95% confidence interval [CI], 0.92-1.12); however, a higher risk was associated with schizophrenia-related disorders (OR, 1.36; 95% CI, 1.02-1.81). Among confirmed cases, mortality risk significantly increased in patients with mental disorders (OR, 1.84, 95% CI, 1.07-3.15).

**Conclusion:** Mental disorders are likely contributing factors of mortality following COVID-19. Although the infection risk did not increase in overall mental disorders, patients with schizophrenia-related disorders were more vulnerable to the infection.

## Introduction

Coronavirus disease 2019 (COVID-19), caused by severe acute respiratory syndrome-coronavirus 2 (SARS-CoV-2), is an ongoing pandemic, affecting 213 countries and yielding more than 11 million cases and 539,000 deaths as of July 9, 2020.(1) Since there is no vaccine and pharmacological therapy currently approved for the disease, it is crucial for government and clinical practitioners to identify vulnerable populations and establish specific strategies for prevention and treatment. So far, older age, obesity, smoking, cardiovascular disease, diabetes, and hypertension have been found as risk factors for COVID-19.(2-6) However, there is no epidemiological evidence on the effect of mental disorders despite of the raised concerns on increased risk of COVID-19 among mentally ill patients.(7, 8)

Patients with mental disorders may be more vulnerable to viral infection than those without, as they have low cognitive ability and are less likely to endeavor for personal protection with little awareness of risk.(8) Moreover, once infected, mentally ill patients may also have a higher risk of severe adverse outcomes for the following reasons: communication difficulties and physicians’ discrimination or negative attitude toward the patients. These may impede receipt of timely medical interventions for COVID-19, resulting in worse prognosis.(9-11) Furthermore, such patients tend to be highly susceptible to stress, and excessive stress caused by restriction of social activities and fear of the epidemic may lead to suppressed immune responses.(12)

Given that mental disorders are highly prevalent worldwide (pooled prevalence estimate of 17.6% across 59 countries),(13) investigating the relation between mental disorders and SARS-CoV-2 infection and severity is important for public health. Here, we assessed the association between mental disorders and the risk of testing positive for SARS-CoV-2, as well as the risk of death and severe events (defined as intensive care unit [ICU] admission, use of mechanical ventilation, and acute respiratory distress syndrome [ARDS]) among confirmed cases using nationwide cohort of patients who received test for SARS-CoV-2.

## Methods

### Study design and data source

We conducted a population-based cohort study using the national health insurance (NHI) claims data from Health Insurance Review & Assessment service (HIRA), linked with the Korea Centers for Disease Control & Prevention (KCDC) data. The data was collected up to May 15, 2020 and contain demographic and clinical information and a 3-year medical history of patients who underwent COVID-19 screening. Information on confirmed cases and those who died from COVID-19 were retrieved from the KCDC data to improve internal validity of database. Clinical information include diagnosis of disease, procedure, inpatient medication orders and prescriptions from all medical institutions in Korea. Disease information was recoded according to the tenth revisions of the International Classification of Diseases (ICD-10). Information on the use of these anonymized data can be obtained through the following website: https://hira-covid19.net/.

### Study population

We constructed two study cohorts: 1) a cohort of patients who received a test for SARS-CoV-2 from December 1, 2019 to May 15, 2020 to investigate the risk of testing positive for SARS-CoV-2, and 2) a sub-cohort of confirmed COVID-19 patients from the first cohort to assess the risk of mortality and severe events following COVID-19. Laboratory confirmation of SARS-CoV-2 was made on the basis of the diagnostic reverse transcription polymerase chain reaction (RT-PCR) test, as recommended by the World Health Organization.(14)

### Assessment of mental disorder

We classified all patients with a diagnosis code for mental and behavioral disorders (ICD-10 codes: F00-F99) within six months prior to the first date of COVID-19 test as patients with mental disorder. Each patients with mental disorder was then matched up to four controls without a diagnosis of mental disorder with the following variables: age (± 2 years), sex, and Charlson Comorbidity Index (CCI). This matching was conducted separately for the overall cohort and sub-cohort.

### Outcome ascertainment

Confirmed COVID-19 cases were ascertained from the KCDC database. To measure severity of COVID-19, we identified two endpoints: death (primary endpoint) and severe events (secondary endpoint). Death cases were also identified using the variable verified by KCDC. We designated 3 health outcomes after infection as severe events, as follows: ICU admission (National Procedure Codes [NPC]: AH190, AH290, AH390, AH110, AH210, AJ001, AJ003, AJ004, AJ005, AJ006, AJ007, AJ008, AJ009, AJ100, AJ200, AJ101, AJ102, AJ201, and AJ202), use of mechanical ventilation (NPC: M0850, M0857, M0858, M0860, M5830, M5850, M5857, M5858, M5860, MM360, and MM400), and ARDS (ICD-10: J80 and P22).

### Potential confounders

The patients’ demographic information (age, sex, type of insurance, and residential area) and clinical baseline characteristics (CCI, comorbidities, and use of co-medications) during 1 year prior to the first date of SARS-CoV-2 test were considered as potential confounders and adjusted in a statistical model. We selected comorbidities, which may be associated with mental disorders and can be risk factors for infection or worse prognosis, as follows: diabetes (E10–E14), hypertension (I10–I15), heart failure (I11.0, I13.0, I13.2, I42, and I50), stroke (I60–64), MI (I21–22, and I252), asthma (J45–J46), COPD (J41–J44), renal disease (I12, I13.1, N03.2–N03.7, N05.2–N05.7, N18, N19, N25.0, Z49.0–Z49.2, Z94.0, and Z99.2), liver disease (B15–19, K70, K71.3–K71.5, K71.7, K72.1, K72.9, K73, K74, and K76), cancer (registration codes: V193,V194, and V027), and pneumonia (J12–J18). Co-medications that are likely to exert confounding effects were included in the study: angiotensin converting enzyme inhibitors, angiotensin receptor blockers, beta-blockers, calcium channel blockers, thiazide diuretics, anticoagulants, anticonvulsants, digoxin, insulin, non-insulin glucose lowering agent, nonsteroidal anti-inflammatory drugs, acetaminophen, and narcotic analgesics.

### Statistical analysis

For the characteristics of the patients, continuous variables are described by mean and standard deviation (SD) and categorical variables are described by frequency and percentage. We calculated the standardized difference to compare the distribution of baseline characteristics between the two groups. Standardized differences greater than 0.1 were considered as imbalances of covariates.

We computed the percentages of patients who were SARS-CoV-2 positive among patients with or without mental disorders, using the cohort of patients who received a SARS-CoV-2 test before and after matching. Odds ratios (ORs) for SARS-CoV-2 infection and their 95% confidence intervals (CIs) were calculated using a logistic regression model for the unmatched cohort and a conditional logistic regression model for the age-, sex-, and CCI-matched cohort, adjusting for type of insurance (health insurance and medical aid), residential area (metropolitan, urban, and rural), comorbidities, and co-medications were included in the models.

In order to examine the effect of mental disorders on severity of COVID-19, we calculated the percentages of patients who died and experienced severe events, among patients with a positive test. We analyzed the data obtained before and after matching, according to the presence or absence of a mental disorder. Likewise, logistic regression and a conditional logistic regression analyses were used for the unmatched and matched cohort, respectively, to estimate ORs for mortality and severe events. We adjusted ORs for the same variables used in the analysis on the risk of a positive test for SARS-CoV-2.

Subgroup analyses were conducted to estimate the risk of SARS-CoV-2 infection and severe COVID-19 among two pre-specified categories of mental disorder based on the ICD-10: schizophrenia, schizotypal and delusional disorders (F20–F29) and mood disorders (F30–F39). These two conditions were chosen as they are considered as severe mental disorders and thus lifestyles and symptoms of patients with these diseases may have different effects on the risk.

As a sensitivity analysis, we calculated the E-value to assess the robustness of the association between mental disorders and COVID-19 mortality to potential unmeasured confounders.

The E-value is the minimum strength of association, which an unmeasured confounder would need to have with both the treatment and outcome on the risk ratio scale, to explain away the observed treatment-outcome association.(15) If the calculated E-value is large, strong unmeasured confounders would be needed to fully explain away an effect estimate.

All analyses were conducted using the SAS statistical software provided by HIRA (SAS Institute Inc, Cary, NC, USA). Two-sided p-values < 0.05 were considered to be statistically significant.

### Ethics approval

The authors assert that all procedures contributing to this work comply with the ethical standards of the relevant national and institutional committees on human experimentation and with the Helsinki Declaration of 1975, as revised in 2008. All procedures involving human subjects/patients were approved by the Institutional Review Board of the Sungkyunkwan University in Korea (IRB number: SKKU-2020-05-012). This observational study was approved with a waiver of informed consent of patients.

## Results

### Characteristics of the study population

A total of 230,565 patients received a laboratory test for SARS-CoV-2 as of May 15, 2020. Among them, 33,653 (14.6%) had mental disorders and 196,912 (85.4%) did not (Figure 1); the mean age of those with and those without mental disorders was 62.4 (SD=21.4) and 44.5 (SD=20.6) years, respectively (Supplementary Table 1). For 24,601 patients with mental disorders, 98,171 controls were matched by age, sex, and CCI. In the matched cohort, the mean age of the two groups was about 55 years and 45.8% were male (Table 1). We identified 7,077 confirmed cases from the first study cohort. Of these patients, 928 (13.1%) were with mental disorders and 6,149 (86.9%) were those without. After matching, there were 743 and 2,865 patients in the mental disorder group and the reference group, respectively. We found that the baseline covariates of the matched cohort were relatively well-balanced compared with those of the unmatched cohort.

**Figure 1.**
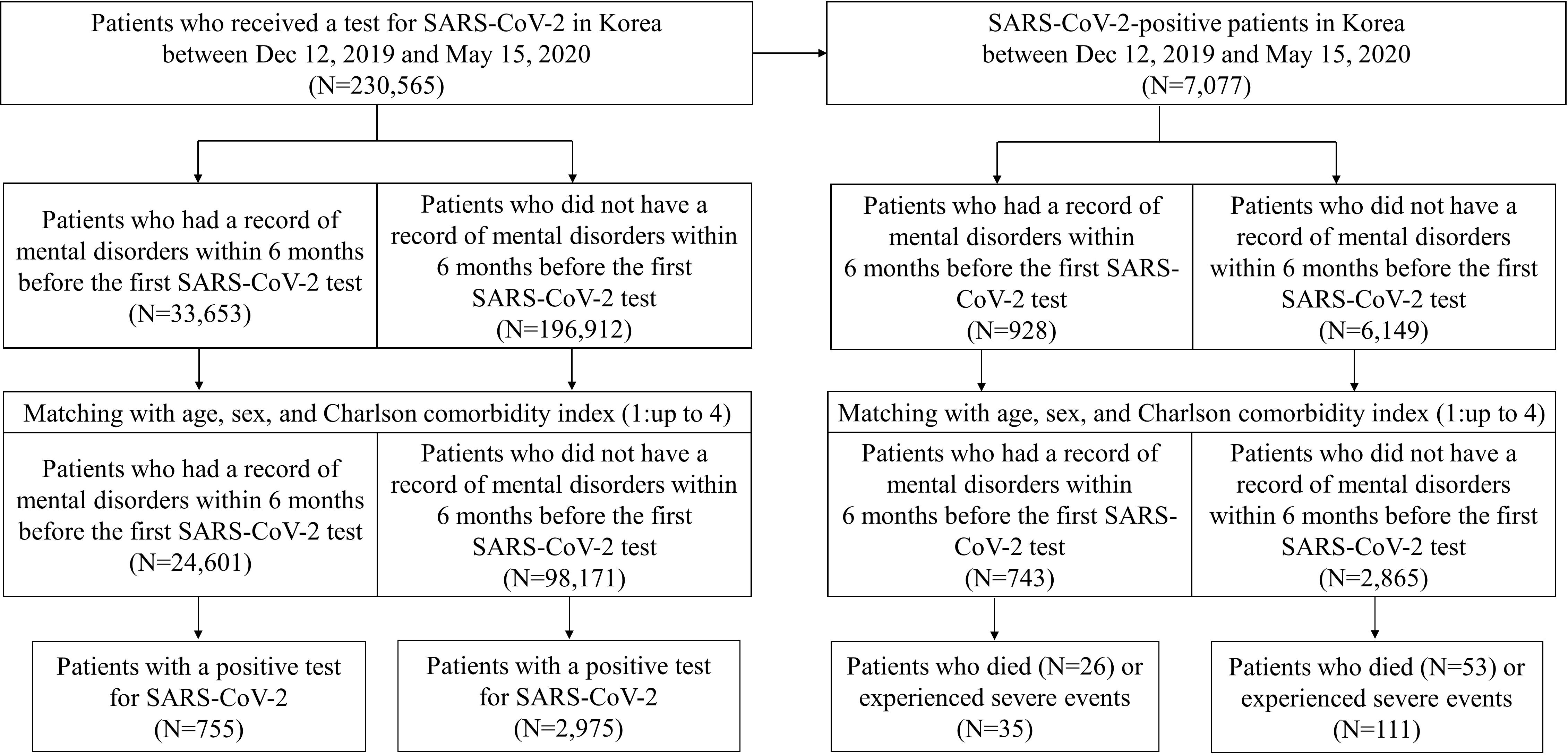
Flow chart of study population selection in the nationwide COVID-19 database from Dec 1, 2019 to May 15, 2020. Abbreviations: COVID-19, coronavirus disease; SARS-CoV-2, severe acute respiratory syndrome coronavirus 2.

**Table 1.** Baseline characteristics of patients with and without mental disorders who underwent tests for SARS-CoV-2 and those who were diagnosed with COVID-19 after age-, sex-, and Charlson Comorbidity Index-matching.

| Characteristics | Patients who underwent tests for SARS-CoV-2<br>N = 122,772 |  |  |  |  | Patients who were diagnosed with COVID-19<br>N = 3,608 |  |  |  |  |
| --- | --- | --- | --- | --- | --- | --- | --- | --- | --- | --- |
|  | With mental disorder |  | Without mental disorder |  | aSD | With mental disorder |  | Without mental disorder |  | aSD |
|  | N = 24,601 (%) |  | N = 98,171 (%) |  |  | N = 743 (%) |  | N = 2,865 (%) |  |  |
| Age (years) (Mean ± SD) | 55.0 | ± 20.2 | 54.8 | ± 20.0 | 0.01 | 56.4 | ± 16.5 | 55.6 | ± 16.3 | 0.05 |
| Male (%) | 11,273 | (45.8) | 44,986 | (45.8) | 0.00 | 317 | (42.7) | 1,211 | (42.3) | 0.01 |
| Residential district |  |  |  |  | 0.08 |  |  |  |  | 0.20 |
| Metropolitan | 6,756 | (27.5) | 29,780 | (30.3) |  | 37 | (5.0) | 247 | (8.6) |  |
| Urban | 5,353 | (21.8) | 22,156 | (22.6) |  | 50 | (6.7) | 284 | (9.9) |  |
| Rural | 12,492 | (50.8) | 46,235 | (47.1) |  | 656 | (88.3) | 2,334 | (81.5) |  |
| Charlson Comorbidity Index (Mean ± SD) | 1.16 | ± 1.53 | 1.21 | ± 1.78 | 0.03 | 0.72 | ± 1.09 | 0.68 | ± 1.07 | 0.04 |
| 0-1 | 16,556 | (67.3) | 66,188 | (67.4) |  | 571 | (76.9) | 2,262 | (79) |  |
| 2 | 4,123 | (16.8) | 16,391 | (16.7) |  | 120 | (16.2) | 428 | (14.9) |  |
| 3+ | 3,922 | (15.9) | 15,592 | (15.9) |  | 52 | (7.0) | 175 | (6.1) |  |
| Comorbidities<sup>a</sup> |  |  |  |  |  |  |  |  |  |  |
| Diabetes mellitus | 4,810 | (19.6) | 17,547 | (17.9) | 0.04 | 140 | (18.8) | 421 | (14.7) | 0.11 |
| Hypertension | 7,557 | (30.7) | 28,132 | (28.7) | 0.05 | 211 | (28.4) | 738 | (25.8) | 0.06 |
| Heart failure | 865 | (3.5) | 4,494 | (4.6) | 0.05 | 15 | (2.0) | 70 | (2.4) | 0.03 |
| Stroke | 1,677 | (6.8) | 4,901 | (5.0) | 0.08 | 36 | (4.8) | 101 | (3.5) | 0.07 |
| Myocardial infarction | 299 | (1.2) | 1,233 | (1.3) | 0.00 | 2 | (0.3) | 24 | (0.8) | 0.08 |
| Asthma | 2,731 | (11.1) | 8,165 | (8.3) | 0.09 | 46 | (6.2) | 150 | (5.2) | 0.04 |
| COPD | 5,737 | (23.3) | 18,624 | (19) | 0.11 | 115 | (15.5) | 376 | (13.1) | 0.07 |
| Renal disease | 1,206 | (4.9) | 4,833 | (4.9) | 0.00 | 4 | (0.5) | 31 | (1.1) | 0.06 |
| Liver disease | 3,332 | (13.5) | 13,469 | (13.7) | 0.01 | 79 | (10.6) | 321 | (11.2) | 0.02 |
| Cancer | 2,636 | (10.7) | 7,861 | (8.0) | 0.09 | 37 | (5.0) | 120 | (4.2) | 0.04 |
| Pneumonia | 2,187 | (8.9) | 14,437 | (14.7) | 0.18 | 22 | (3.0) | 151 | (5.3) | 0.12 |
| Concomitant medications<sup>§</sup> |  |  |  |  |  |  |  |  |  |  |
| ACE inhibitors | 336 | (1.4) | 1,440 | (1.5) | 0.01 | 10 | (1.3) | 33 | (1.2) | 0.02 |
| Angiotensin II receptor blockers | 6,219 | (25.3) | 23,637 | (24.1) | 0.03 | 161 | (21.7) | 603 | (21.0) | 0.02 |
| β-blockers | 7,935 | (32.3) | 14,199 | (14.5) | 0.43 | 202 | (27.2) | 284 | (9.9) | 0.46 |
| Calcium channel blockers | 6,294 | (25.6) | 23,018 | (23.4) | 0.05 | 153 | (20.6) | 500 | (17.5) | 0.08 |
| Thiazide diuretics | 2,357 | (9.6) | 7,908 | (8.1) | 0.05 | 61 | (8.2) | 215 | (7.5) | 0.03 |
| Anticoagulants | 9,322 | (37.9) | 30,759 | (31.3) | 0.14 | 189 | (25.4) | 595 | (20.8) | 0.11 |
| Anticonvulsants | 6,793 | (27.6) | 12,527 | (12.8) | 0.38 | 166 | (22.3) | 198 | (6.9) | 0.45 |
| Digoxin | 268 | (1.1) | 1,348 | (1.4) | 0.03 | 4 | (0.5) | 15 | (0.5) | 0.00 |
| Insulin | 1,973 | (8.0) | 7,578 | (7.7) | 0.01 | 36 | (4.8) | 70 | (2.4) | 0.13 |
| Non-insulin glucose lowering agents | 3,970 | (16.1) | 15,647 | (15.9) | 0.01 | 127 | (17.1) | 380 | (13.3) | 0.11 |
| NSAIDs | 21,397 | (87.0) | 80,933 | (82.4) | 0.13 | 587 | (79.0) | 2,313 | (80.7) | 0.04 |
| Acetaminophen | 18,939 | (77.0) | 66,974 | (68.2) | 0.20 | 513 | (69.0) | 1,818 | (63.5) | 0.12 |
| Narcotic analgesics | 15,440 | (62.8) | 54,336 | (55.3) | 0.15 | 429 | (57.7) | 1,504 | (52.5) | 0.11 |

SARS-CoV-2, severe acute respiratory syndrome coronavirus 2; COVID-19, coronavirus disease; ACE, angiotensin converting enzyme inhibitors; aSD, absolute standardized difference; COPD, chronic obstructive pulmonary disease; NSAIDs, non-steroidal anti-inflammatory drugs; SD, standard deviation.
<sup>a</sup> Comorbidities and concomitant medications were assessed within a year before the cohort entry date.

**Table 2.** Risk of a positive test for SARS-CoV-2 and severe outcomes following COVID-19

|  | No. of patients | No. of events | Percentage of events<sup>a</sup> (%) | Odds ratio |  |
| --- | --- | --- | --- | --- | --- |
|  |  |  |  | Crude (95% CI) | Adjusted<sup>b</sup> (95% CI) |
| **SARS-CoV-2-positive patients among those who received a SARS-CoV-2 test** |  |  |  |  |  |
| *Before matching* |  |  |  |  |  |
| With mental disorder | 33,653 | 928 | 2.76 | 0.88 (0.82–0.94) | 0.95 (0.87–1.03) |
| Without mental disorder | 196,921 | 6,149 | 3.12 | 1.00 (ref) | 1.00 (ref) |
| *After matching<sup>c</sup>* |  |  |  |  |  |
| With mental disorder | 24,601 | 755 | 3.07 | 1.01 (0.93–1.10) | 1.02 (0.92–1.12) |
| Without mental disorder | 98,171 | 2,975 | 3.03 | 1.00 (ref) | 1.00 (ref) |
| **Patients experienced severe outcomes following COVID-19 among confirmed patients** |  |  |  |  |  |
| *Before matching* |  |  |  |  |  |
| Death (primary) |  |  |  |  |  |
| With mental disorder | 928 | 56 | 6.03 | 7.12 (4.87–10.39) | 3.93 (2.57–6.03) |
| Without mental disorder | 6,149 | 55 | 0.89 | 1.00 (ref) | 1.00 (ref) |
| Severe events<sup>d</sup> (secondary) |  |  |  |  |  |
| With mental disorder | 928 | 44 | 4.74 | 2.31 (1.63–3.27) | 1.47 (0.99–2.19) |
| Without mental disorder | 6,149 | 130 | 2.11 | 1.00 (ref) | 1.00 (ref) |
| *After matching<sup>c</sup>* |  |  |  |  |  |
| Death (primary) |  |  |  |  |  |
| With mental disorder | 743 | 26 | 3.50 | 1.92 (1.20–3.10) | 1.84 (1.07–3.15) |
| Without mental disorder | 2,865 | 53 | 1.85 | 1.00 (ref) | 1.00 (ref) |
| Severe events<sup>d</sup> (secondary) |  |  |  |  |  |
| With mental disorder | 743 | 35 | 4.71 | 1.23 (0.83–1.81) | 1.17 (0.76–1.81) |
| Without mental disorder | 2,865 | 111 | 3.87 | 1.00 (ref) | 1.00 (ref) |

CI, confidence interval; SARS-CoV-2, severe acute respiratory syndrome coronavirus 2; COVID-19, coronavirus disease
<sup>a</sup> No. of events/no. of patients ×100
<sup>b</sup> Adjusted for type of insurance, residential area, comorbidities, and co-medications.
<sup>c</sup> The cohort matched with age, sex, and Charlson Comorbidity Index.
<sup>d</sup> Defined as intensive care unit (ICU) admission, use of mechanical ventilation, and acute respiratory distress syndrome (ARDS).
in patients with mental disorders compared to those without mental disorders.

### The risk of positive test for SARS-Co V-2 in patients with mental disorders

In the unmatched cohort, there were 2.76% and 3.12% COVID-19 patients with and without mental disorder, respectively. The risk of a SARS-CoV-2-positive test in patients with mental disorders was 0.95 (95% CI, 0.87–1.03). After matching, the percentage of SARS-CoV-2-positive patients in both groups was approximately 3.0%, and the fully adjusted OR was 1.02 (95% CI, 0.92–1.12).

### The risk of severe COVID-19 in patients with mental disorders

Among the 928 patients with mental disorders, 56 (6.03%) died and 44 (4.74%) experienced severe events. The percentages of patients who died and had severe events were 0.89% and 2.11%, respectively, in patients without mental disorders. The ORs of death and severe events were 3.93 (95% CI, 2.57–6.03) and 1.47 (95% CI, 099–2·19), respectively. In the matched cohort, 26 (3.50%) of 743 patients with mental disorders and 53 (1.85%) of 2,865 patients without mental disorders died. Compared with patients without mental disorders, the risk of death was increased in multivariate analysis (OR, 1.84; 95% CI, 1.07–3.15). The E-value obtained from the estimate was 3.08. The percentage of patients who had severe events was 4.71% (35/743) in the mental disorders group and 3.87% (111/2,865) in the reference group. The association between mental disorders and severe events was not statistically significant (OR, 1.17; 95% CI, 0.76–1.81).

### Subgroup analysis

When we repeated analysis by restricting to those with schizophrenia, schizotypal and delusional disorders and mood disorders, the adjusted ORs were 1.36 (95% CI, 1.02–1.81) in patients with schizophrenia and schizotypal and delusional disorders, and 0.78 (95% CI, 0.68–0.90) in patients with mood disorders (Table 3). The risk of mortality and severe events after SARS-CoV-2 infection were higher in patients with schizophrenia and schizotypal and delusional disorders, but the effect was not statistically significant (death: adjusted OR, 2.71; 95% CI, 0.11–66.17, severe events: adjusted OR, 2.69; 95% CI, 0.62–11.67). In the cohort with mood disorders, there was a significantly increased risk of death (adjusted OR, 3.57; 95% CI, 1.36–9.38), but the risk of severe events was not significant (adjusted OR, 1.48; 95% CI 0.70–3.13).

**Table 3.**
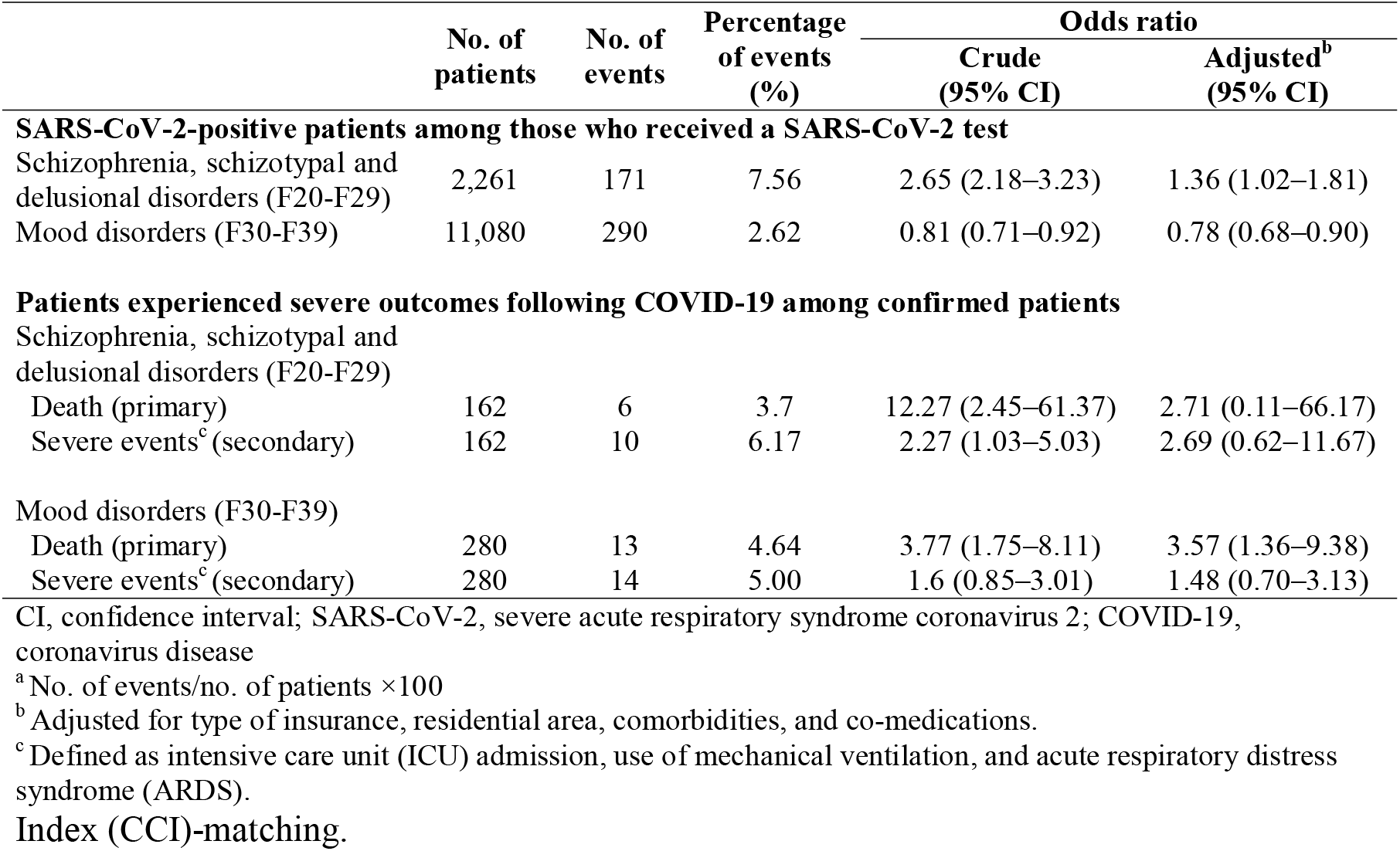
Risk of a positive test for SARS-CoV-2 and severe outcomes following COVID-19 in patients with schizophrenia, schizotypal and delusional disorders, and mood disorders compared to those without mental disorders, after age-, sex-, and Charlson Comorbidity

### Patient and Public Involvement

Patients or the public could not be involved in the design, or conduct, or reporting, or dissemination plans of our research since we used de-identified patient data.

## Discussion

Evidence from this nationwide cohort study suggests that mental disorders did not increase the risk of SARS-CoV-2 infection but were likely to increase mortality risk upon infection. With regard to severe events, we did not find an increased risk among SARS-CoV-2-positive patients. The risk of a SARS-CoV-2-positive test was higher in patients who had schizophrenia and schizotypal and delusional disorders. We also found that the mortality risk was significantly higher in patients with mood disorders.

Our data showed an increased mortality risk following COVID-19 among patients with mental disorders. This finding is in line with a study, which has found a significant relation between mental disorders and cause-specific mortality. (16) The positive association may not be attributed solely to the mental disorder and may be multifactorial. Since mental illness are generally associated with unhealthy lifestyle (e.g. smocking and alcohol consumption) and a low socioeconomic status,(17–19) these factors may also contribute to a worse prognosis of COVID-19. In addition, the use of antipsychotics may increase the risk of death or severe events. Some antipsychotic medications are known to have immunomodulatory effects.(20) Chlorpromazine and clozapine can affect the production of cytokines, resulting in suppression of immune response. Furthermore, health inequality in people with mental illness have been suggested as an important contributor of poor physical health outcomes.(10, 11, 21) Stigma and discrimination toward mental illness and poor communication skills of patients with mental disorders may hamper proper and timely provision of medical intervention for COVID-19 and therefore caused severe health outcomes.

Previous studies found a positive association between severe mental illness and microbial or viral infection (pneumonia, human immunodeficiency virus, hepatitis B, and hepatitis C).(22, 23) In our study, patients with schizophrenia-related disorders were at risk of SARS-CoV-2 infection although no association was observed among patients with overall mental disorders. Patients with schizophrenia-related disorders may be more vulnerable to SARS-CoV-2 infection than those with other mental disorders since they are likely to live in shared accommodation such as a psychiatric hospital. Unlike patients in general hospitals, those in psychiatric hospitals commonly participate in group activities, creating conditions favorable for virus transmission. Additionally, teaching such patients to follow personal control measures would be difficult due to their impaired cognitive ability. (24) By contrast, we found that the infection risk decreased in patients with mood disorders. One possible explanation may be that patients with mood disorders would engage in fewer social activities than would healthy individuals, reducing the chance of exposure to the virus. Indeed, COVID-19 has spread among young and healthy people due to the highly contagious nature of the virus, though the fatality rate is low.(25, 26)

To the best of our knowledge, there are no published studies investigating the effect of mental disorders as a risk factor for susceptibility and severity of COVID-19 so far. We also used the nationwide COVID-19 database that include medical histories of overall COVID-19 cases in Korea, which strengthens generalizability of our results. Moreover, we have enhanced the internal validity by using information on confirmed COVID-19 cases and death cases obtained from the KCDC. The Korean government has managed confirmed and suspected COVID-19 cases strictly. According to the government’s response system, all inbound travelers are monitored and tested if they present with fever or respiratory symptoms. If a patient tests positive for SARS-CoV-2, all primary patient contacts identified by epidemiological investigation receive a test if they exhibit symptoms during a 14-day self-quarantine period.(27) COVID-19 patients classified moderate, severe, and extremely severe cases are hospitalized and managed by the government.(28) Therefore, underestimation of confirmed and death cases would be trivial.

This study has some limitations. First, covariates regarding lifestyle (smoking status and alcohol consumption) and socioeconomic status (education and income level) of patients were not included in the analytic model. The NHI database was constructed on the basis of claims data and thus these variables were not available. Although unmeasured confounders are known to be associated with worse outcomes after SARS-CoV-2 infection, the result from the sensitivity analysis suggests that an unmeasured confounder associated with both mental disorders and death by a risk ratio of 3.08-fold each is required to explain away the observed OR of 1.84. Hence, our findings would be robust unless there is an unmeasured confounder of such magnitude. Second, we identified severe events based on diagnostic and procedure codes, which were recorded for administrative purpose; thus, there was potential for misclassification of outcomes. However, a validation study comparing the claims database and inpatients’ medical records of hospitals reported that the overall agreement of diagnosis was 82.0%.(29) Procedure codes, which are directly related to payment from the NHI, are also likely to be highly valid. Third, although we utilized the database covering overall COVID-19 patients in Korea, the number of patients with mental disorders was not sufficient to evaluate the risk of death and severe events by subgroup analysis based on more subdivided disease type such as depression, anxiety, and dementia. Further studies would be needed to determine whether patients with certain specific diseases have a higher risk of SARS-CoV-2 infection and severe COVID-19.

In summary, our results suggest that mental disorders were likely to increase the risk of death following COVID-19. We found no association between overall mental disorders and the risk of SARS-CoV-2 infection among patients who were tested; however, the risk was significantly higher in patients with schizophrenia-related disorders. Psychiatrists should inform patients and their caregivers about the risk of SARS-CoV-2 infection and guide them to comply with preventive measures. Furthermore, clinicians and healthcare policy makers would need to pay more attention to patients with mental disorders during the COVID-19 pandemic and establish preventive strategies from them.

## Data Availability

The data that support the findings of this study are available from Health Insurance Review & Assessment service (HIRA) in South Korea. Restrictions apply to the availability of these data, which were used under licence for this study. Data access is available at https://hira-covid19.net/ with the permission of HIRA.

## Acknowledgments

The authors appreciate healthcare professionals dedicated to treating COVID-19 patients in Korea, and the Ministry of Health and Welfare, the Health Insurance Review & Assessment Service of South Korea for sharing invaluable national health insurance claims data in a prompt manner. We especially thank Do-Yeon Cho of the Health Insurance Review & Assessment Service for executing the analysis software. We also thank Ju Hwan Kim of Sungkyunkwan University and Editage (www.editage.co.kr) for English language editing.

## Author Contribution

HLJ and JYS designed the study. SHP analyzed the data. HLJ, SHP, and JSK interpreted the data. HLJ wrote the first draft. JSK and JYS critically revised the draft. All authors approved the final manuscript. A guarantor and corresponding author JYS had full access to the data, takes responsibility for the integrity of the data, and controlled the decision to publish. The corresponding author attests that all listed authors meet authorship criteria and that no others meeting the criteria have been omitted.

